# Developing an SNR-efficient tensor-valued diffusion encoding protocol for studying brain microstructural changes in neurodegeneration

**DOI:** 10.64898/2026.09.16.26363267

**Authors:** Erpeng Dai, Xuetong Zhou, S Shailja, Martin K Schneider, Molly A Millar, Michael Zeineh, Carl-Fredrik Westin, Jennifer A McNab

## Abstract

Tensor-valued diffusion encoding (TDE) is an emerging diffusion MRI technique that uses advanced diffusion-encoding waveforms and modeling to provide enhanced specificity to brain microstructure compared with conventional Stejskal-Tanner pulsed-gradient encoding. However, the diffusion encoding duration to achieve the same b-value is substantially increased in TDE, resulting in relatively low SNR and spatial resolution (>2 mm isotropic). In this study, we developed an SNR-efficient, high-resolution TDE protocol with 1.8-mm isotropic resolution and clinically feasible scan time and evaluated its reproducibility and sensitivity to age-related microstructural differences. Eleven cognitively normal older adults (5F/6M, 62-71 years) and seven younger adults (3F/4M, 22-31 years) were scanned at 3T using an in-house TDE sequence incorporating an SNR-efficient multi-band multi-shot EPI readout and reconstruction. Quantitative diffusion metrics, including mean diffusivity (MD), fractional anisotropy (FA), microscopic FA (μFA), anisotropic mean kurtosis (MK_A_), isotropic mean kurtosis (MK_I_), and total mean kurtosis (MK_T_), were evaluated in the bilateral temporal parts of the cingulum bundles (CBT), bilateral fornix (FX), and global white matter (WM). Older participants underwent two scans to assess reproducibility using intraclass correlation coefficients (ICCs) and Bland-Altman analysis, and diffusion metrics were compared between younger and older groups using Wilcoxon signed-rank tests with false discovery rate (FDR) correction. All metrics except MK_I_ demonstrate moderate-to-excellent reproducibility across all WM regions (ICCs≥0.64). Exploratory comparisons show age-related differences in FA, μFA, and MK_A_ with *p*<0.05 before FDR correction, particularly in the fornix, although none remains significant after correction. These findings demonstrate the feasibility of SNR-efficient TDE at 1.8-mm isotropic resolution with approximately 70-mm brain coverage in approximately 12 minutes and establish its reproducibility for high-resolution brain microstructural imaging.

**Key Points:**

1. An SNR-efficient tensor-valued diffusion encoding (TDE) protocol was developed with 1.8-mm isotropic resolution.
2. The high-resolution TDE protocol achieved ∼70-mm brain coverage within a clinically feasible scan time of ∼12 minutes.
3. The developed TDE protocol was evaluated in two white matter bundles closely associated with neurodegeneration and global white matter.
4. All quantitative diffusion metrics except MK_I_ demonstrate moderate-to-excellent scan-rescan reproducibility in across white matter ROIs (ICC≥0.64).
5. Exploratory comparisons show age-related differences in FA, μFA, and MK_A_ before FDR correction, particularly in the fornix.

## 1 Introduction

Diffusion MRI is a promising technique for detecting brain microstructural changes associated with neurodegeneration in normal aging and disease (1). Widespread decrease in fractional anisotropy (FA) and increase in mean diffusivity (MD) have been observed during normal aging (2–4). Moreover, the extent of FA and MD changes is further aggravated in neurodegenerative disorders, such as Alzheimer’s disease (AD) (5–8). AD-specific studies have also revealed that diffusion property changes are closely associated with AD pathology, clinical progression, and cognitive impairments (9–11).

Numerous diffusion MRI protocols have been established for neurodegeneration studies. For instance, a basic single-shell (*b*=1 ms/μm^2^) diffusion tensor imaging (DTI) protocol and an advanced multi-shell (*b*=0.5, 1, and 2 ms/μm^2^) high angular resolution diffusion imaging (HARDI) protocol have been built for the Alzheimer’s Disease Neuroimaging Initiative (ADNI) (12,13). With higher b-values, the HARDI protocol enables the use of advanced diffusion models, such as diffusion kurtosis imaging (DKI) (14), to elucidate microstructural changes beyond FA and MD. However, despite more and higher b-values than DTI, HARDI still uses the standard Stejskal-Tanner pulsed-gradient spin-echo (PGSE) sequence, i.e., a pair of identical trapezoid diffusion-encoding gradients before and after the 180° refocusing pulse (15). The PGSE encoding is only sensitive to the average effects of diffusion motion across the imaging voxel and thus provides limited specificity in characterizing brain microstructural changes.

Tensor-valued diffusion encoding (TDE) is a new diffusion encoding framework in which diffusion encoding is characterized by a full B-tensor (a 3×3 matrix), rather than a single-direction b-vector (a 3×1 vector) (16). Within this framework, the standard PGSE sequence corresponds to a B-tensor with only one nonzero principal direction and is therefore referred to as linear tensor encoding (LTE). Similarly, higher-order diffusion encoding waveforms can be defined, including planar tensor encoding (PTE) and spherical tensor encoding (STE), which correspond to a planar and spherical B-tensor shape, respectively. More details of the TDE framework, along with example LTE, PTE, and STE gradient waveforms, q-space trajectories, and associated B-tensor shapes, can be found in Refs. (16–18). In practice, the implementation of TDE comprises LTE combined with either PTE or STE, or both.

Compared with DTI and DKI, a notable advantage of TDE is the improved specificity for brain microstructure detection. Specifically, in addition to uncovering the average diffusion effects with FA and MD, TDE can better distinguish between variances in diffusion effects, such as diffusivity, orientation, and microscopic diffusion anisotropy (16,19). For instance, the microscopic FA (μFA) parameter derived from TDE is sensitive to microscopic diffusion anisotropy but insensitive to only to diffusion orientation dispersion, whereas FA derived from DTI reflects a mixed effect of microscopic diffusion anisotropy, diffusivity variations, and orientation dispersion. TDE has recently demonstrated promising clinical utility for estimating microscopic anisotropy and tissue heterogeneity in brain tumors within clinically feasible acquisition times (20,21).

Despite its great potential for more specific characterization of brain microstructure, the application of TDE in neurodegenerative research remains limited. A primary reason is the SNR challenge for imaging the neurodegeneration-associated brain regions, such as the medial temporal lobe (MTL) (22). The MTL lies relatively deep within the brain and comprises multiple important white matter (WM) and gray matter (GM) structure that support cognitive function, such as the temporal part of the cingulum bundle (CBT), fornix (FX), and hippocampus (23). The SNR performance is lower in deep brain regions than in the superficial cortex, a limitation that can be further exacerbated when high spatial resolution is required to resolve fine-scale WM bundles, such as the FX (24). Second, high-order diffusion encoding waveforms (i.e., PTE and STE) can substantially prolong the diffusion-encoding duration to achieve the same b-value as LTE (17). The prolonged diffusion-encoding duration further results in a longer echo time (TE), more T_2_ decay, and a lower SNR. Lastly, high b-values (*b*≥2 ms/μm^2^) are needed for advanced diffusion modeling in TDE, which not only increases the diffusion encoding duration but also induces more diffusion decay, further exacerbating the SNR challenge.

Substantial efforts have been invested in improving the SNR efficiency of diffusion MRI, from hardware (25–27), diffusion encoding (17,28), and readout (29–32). Specifically, for TDE, asymmetric diffusion-encoding waveforms have been developed to reduce the diffusion-encoding duration (or equivalently, improve the diffusion-encoding efficiency) of PTE and STE (17). From the image readout aspect, a super-resolution reconstruction method based on rotating-view acquisitions has been proposed to retain sufficient SNR at high spatial resolution and high b-values (32). With this super-resolution technique, 1.6 mm isotropic high resolution was reconstructed from an acquisition with a lower through-plane resolution of 7.2 mm. However, implementing super-resolution required rotating each low-resolution acquisition multiple times, resulting in an acquisition time of 33.6 s per diffusion volume and limiting its clinical applicability.

The primary objective of this work is to develop a high-resolution (1.8 mm isotropic) TDE protocol for imaging the MTL within clinically feasible scan times. As an initial validation of the TDE protocol, we evaluate the scan-rescan reproducibility of quantitative TDE metrics in cognitively normal older adults, with the goal of establishing the protocol for future studies of neurodegeneration. Additionally, as a proof-of-concept application, we examine age-related differences in TDE-derived quantitative metrics between cognitively normal younger and older adults.

## 2 Theory and Methods

### 2.1 Sequence design

This study was conducted on a 3T GE scanner equipped with a UHP gradient coil (G_max_=100 mT/m and SR_max_=200 T/m/s) (GE Healthcare, Waukesha, WI), and a 32-channel phased-array receive coil (Nova Medical, USA). An in-house TDE sequence was developed by jointly using LTE and STE waveforms (Figure 1). For STE, Maxwell-compensated asymmetric gradient waveforms were designed using the MATLAB numerical optimization toolbox (Figure 1A) to improve diffusion-encoding efficiency while mitigating concomitant gradient effects (33–35). The optimization constraints included: Maxwell index threshold=100 (mT/m)^2^ms, total duration=72.64 ms, G’_max_=85 mT/m, and SR’_max_=134 T/m/s. The G’_max_ was slightly derated from the nominal maximum gradient amplitude of the gradient coil (G_max_=100 mT/m) to achieve a higher duty cycle. For LTE, a pair of symmetric trapezoid gradient waveforms with the same total duration as the optimized STE waveform were used (Figure 1B).

**Figure 1:**
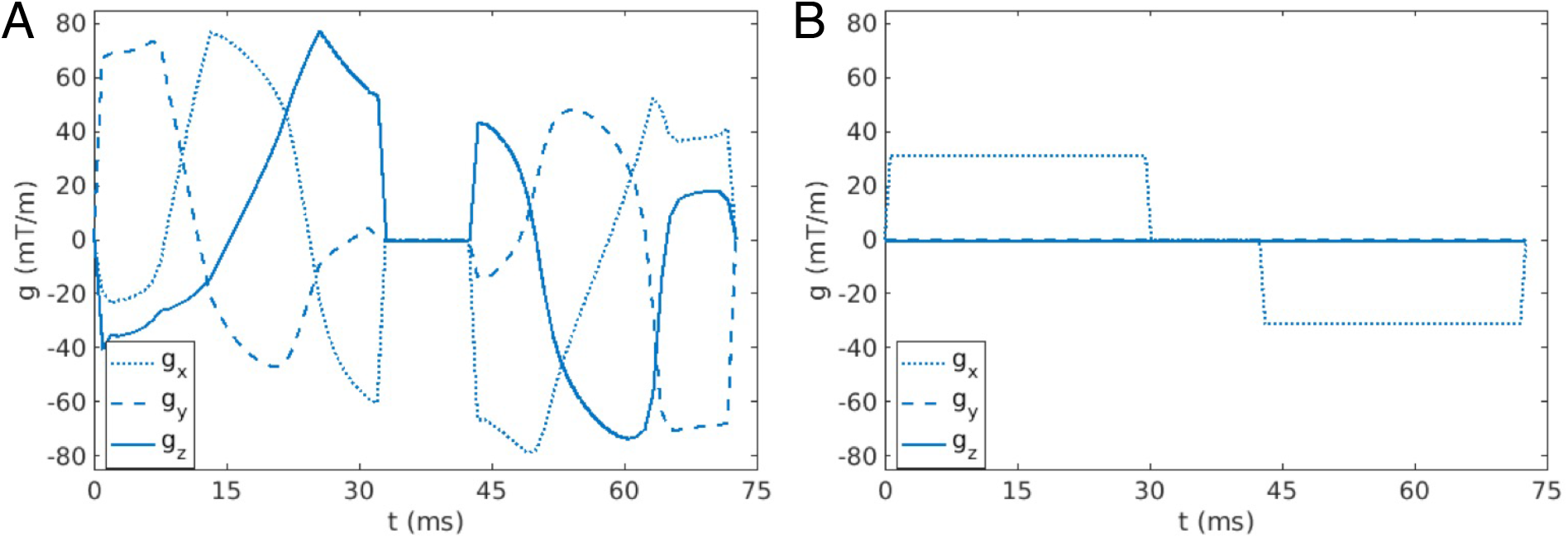
A customized tensor-valued diffusion encoding (TDE) sequence is developed by jointly using linear and spherical tensor-valued diffusion encoding (LTE and STE). (**A**) A Maxwell-compensated asymmetric gradient waveforms for STE. (**B**) A pair of symmetric gradient waveforms with the same diffusion encoding duration for LTE.

### 2.2 Data acquisition and reconstruction

All human MRI scans were approved by the local Institutional Review Board, and written consent was obtained from each subject before the scan. Eleven cognitively normal older subjects (5F/6M, 62∼71 y/o) and seven cognitively normal younger subjects (3F/4M, 22∼31 y/o) were scanned. Each older subject was scanned twice (“older-1” and “older-2”, with a time interval of 6∼18 days between the two scans) to evaluate the scan-rescan reproducibility of the developed TDE protocol, while each younger subject was scanned only once (“younger”).

The imaging parameters of the TDE protocol were: FOV=220×220 mm^2^, 38 axial slices, voxel size=1.8 mm isotropic, TE=81 ms, TR=3.3 s, partial-Fourier (PF) factor=0.63, total scan time (T_acq_)=11min52s. The diffusion encoding scheme of STE included *b*=0.1 (6 averages), 0.7 (6 averages), 1.4 (15 averages), and 2.0 ms/μm^2^ (30 averages), while that of LTE was *b*=0.1 (3 directions), 0.7 (3 directions), 1.4 (12 directions), and 2.0 ms/μm^2^ (24 directions). Six *b*=0 volumes (“blip up”) were evenly spaced between the diffusion volumes, and an additional *b*=0 volume was acquired with reversed phase-encoding polarity (“blip down”) for distortion correction. A multi-band multi-shot (MB-MS) EPI sequence was used for data readout, with MB=2, N_shot_=2, and controlled aliasing in parallel imaging (CAIPI) with FOV/4 shift (36). The q-space sampling was interleaved between different diffusion volumes to distribute energy consumption and heating more evenly over time and to reduce potential bias from system drifts.

A multi-shell HARDI diffusion scan was conducted on each subject to track the CBT and FX, using GE’s product single-shot EPI (ss-EPI) sequence. The detailed scan parameters were: FOV=220×220 mm^2^, 46 axial slices, voxel size=1.5 mm isotropic, MB=2, SENSE=2, CAIPI with FOV/4 shift, TE=60 ms, TR=4.2 s, PF=0.61, T_acq_=9min35s. The diffusion encoding scheme included *b*=1.0 (65 directions) and 2.0 ms/μm^2^ (65 directions). Five “blip-up” *b*=0 volumes were evenly interspaced between the diffusion volumes, and an extra “blip-down” *b*=0 volume was acquired for distortion correction.

A T1-weighted (T1w) MPRAGE scan was also conducted for each subject. The detailed scan parameters were: TR=1924 ms, TE=2.75 ms, inversion time (TI)=900 ms, flip angle=8°, FOV=220×220×160 mm^3^ (sagittal view), voxel size=1.0 mm isotropic, T_acq_=3min55s.

The T1w-MPRAGE and HARDI data were reconstructed online. The TDE data were reconstructed offline using MATLAB 2024b (Mathworks, Natick, MA, USA). The N/2 Nyquist ghost artifact and ramp sampling artifact were corrected using GE’s Orchestra offline reconstruction pipeline. The corrected k-space data were compressed to 12 channels using coil compression (37), and subsequently reconstructed jointly using structured low-rank constraints and explicit phase mapping, as described in Ref. (36).

### 2.3 Image processing and analysis

The image processing and analysis pipeline was summarized in Figure 2 and detailed below.

**Figure 2:**
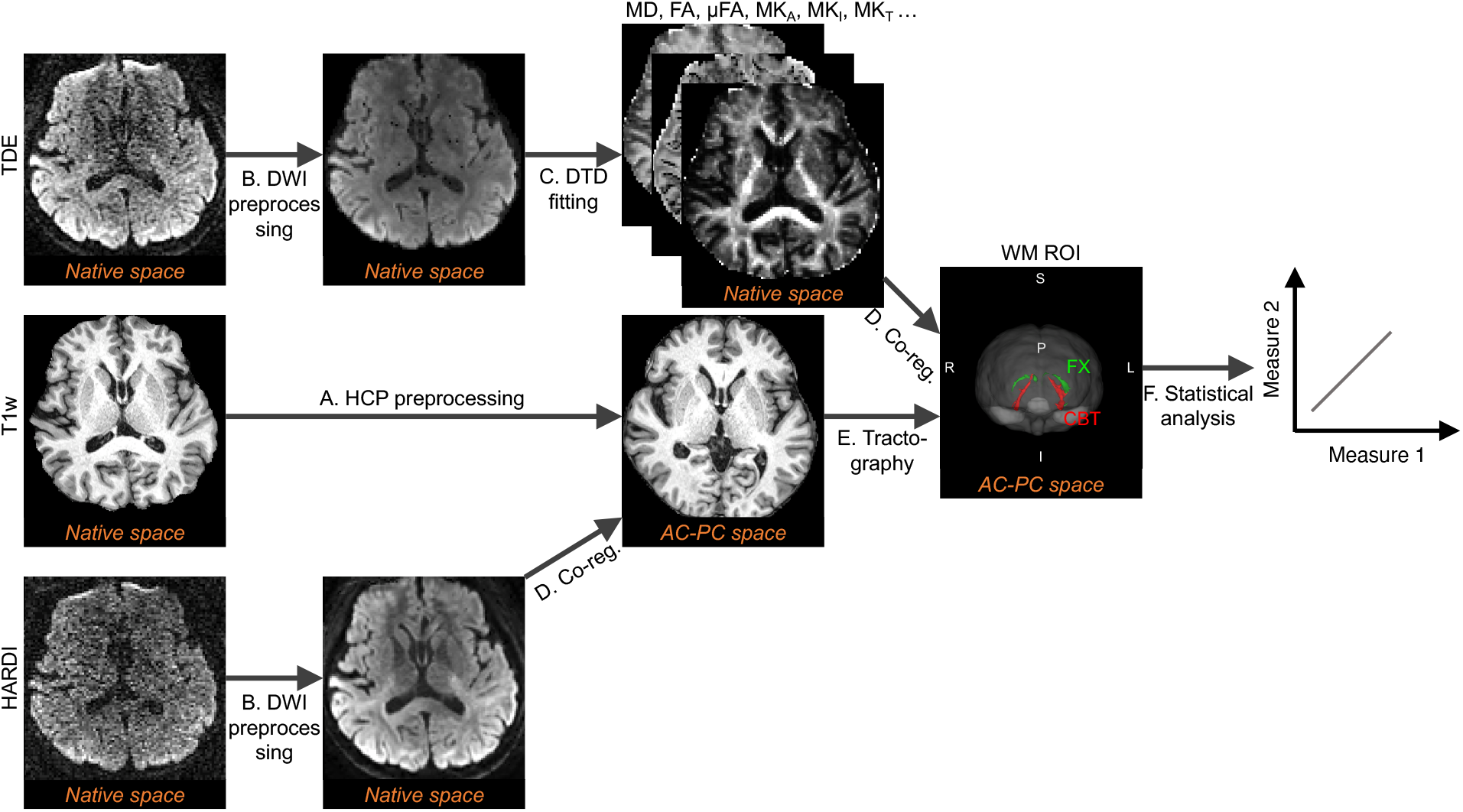
Image processing and analysis pipeline. (A) Converting the T1w-MPRAGE images into the subject’s AC-PC space) using the HCP preprocessing pipeline. (B) Preprocessing of diffusion-weighted images (DWIs) in the native diffusion space. (C) Diffusion fitting in the native diffusion space. (D) Co-registering preprocessed DWIs and quantitative diffusion parameter maps to the AC-PC space. (E) The generated binary white matter ROIs from diffusion tractography with the HARDI data in the AC-PC space. (F) Statistical analysis in the AC-PC space. Abbreviations: MD: mean diffusivity; FA: fractional anisotropy; μFA: microscopic fractional anisotropy; MK_A_: anisotropic mean kurtosis; MK_I_: isotropic mean kurtosis; MK_T_: total mean kurtosis (MK_A_ + MK_I_); TDE: tensor-valued diffusion encoding; HARDI: high angular resolution diffusion imaging; DTD: diffusion tensor distribution; DKI: diffusion kurtosis imaging; HCP: human connectome project; DWI: diffusion-weighted images. The space information (“native” or “AC-PC”) is marked under images at each stage.

#### 2.3.1 Preprocessing

The T1w-MPRAGE images were preprocessed using the Human Connectome Project (HCP) preprocessing pipeline (38) to generate the T1w image in the subject’s AC-PC space (“T1 AC-PC”) (Figure 2A) and the non-linear transformation information between the T1 AC-PC space and the Montreal Neurological Institute (MNI) space.

The preprocessing of the diffusion-weighted images (DWIs) (Figure 2B) included Marchenko-Pastur Principal Component Analysis (MPPCA) denoising (39) and correction of residual Gibbs ringing artifacts (40) using the DESIGNER toolbox (41). The susceptibility-induced distortion artifacts were then corrected using FSL TOPUP on the *b*=0 images of each dataset (42). For the HARDI data, image distortions from eddy currents and motion artifacts were corrected using FSL EDDY (43). For the TDE data, since STE with arbitrary waveforms was used, the induced eddy currents and their influences on distortion are different from those from LTE. Image distortions caused by eddy currents and motion artifacts were corrected using ElastiX (44).

#### 2.3.2 Diffusion parameter fitting

Quantitative diffusion metrics, including MD, FA, μFA, anisotropic mean kurtosis (MK_A_), isotropic MK (MK_I_), and total MK (MK_T_), were fitted from the preprocessed TDE data as in Ref. (21). Specifically, all volumes were first smoothed by a 3D Gaussian kernel with a standard deviation (SD) of 0.4 voxels. The diffusion tensor distribution covariance model was then used for the fitting, using the MATLAB MD-DMRI toolbox (Figure 2C) (45), A brief review and comparison of the derived diffusion metrics are given below. The difference between FA and μFA was reviewed in the Introduction section, i.e., μFA was sensitive to microscopic diffusion anisotropy but insensitive to orientation dispersion, while FA was a mixed effect of microscopic diffusion anisotropic and orientation dispersion. MK_A_ reflected the portion of kurtosis caused by microscopic diffusion anisotropy (“variance in shape”), whereas MK_I_ was associated with the portion of kurtosis due to heterogeneity in isotropic diffusivities (“variance in magnitude”). The sum of MK_I_ and MK_A_ yielded the total mean kurtosis (MK_T_), which was similar but not identical to MK obtained from DKI, as they came from different diffusion encoding mechanisms and modelling assumptions (16). While both μFA and MK_A_ both reflected microscopic anisotropy, μFA directly quantified its magnitude, and MK_A_ quantified how the microscopic anisotropy contributes to the non-Gaussian effects in diffusion, i.e., kurtosis.

#### 2.3.3 Co-registration to the T1 AC-PC space

To enable a direct scan-rescan comparison for the older subjects, either the preprocessed DWIs or the fitted diffusion parameter maps were co-registered to the T1 AC-PC space, using FreeSurfer BBREGISTER (46) (Figure 2D).

#### 2.3.4 Segmentation of WM ROIs

Diffusion tractography was conducted in each subject’s T1 AC-PC space by co-registering the HARDI data and rotating the b-vector to the T1 AC-PC space. Voxel-wise fiber orientation distributions were estimated using GPU-accelerated BEDPOSTX in FSL (47). The bilateral CBT and FX bundles were tracked in the T1 AC-PC space using GPU-accelerated PROBTRACKX in FSL (48). The seed, target, exclusion, and/or stop masks were generated by co-registering the predefined template masks in the MNI space from FSL XTRACT (49) to the T1 AC-PC space, using the non-linear transformation information generated from the above HCP preprocessing (Figure 2A). The normalized fiber probability maps of CBT and FX were further binarized by: FA≥0.2, MD≤1.5 μm^2^/ms, and a percentage threshold of the normalized fiber probability of 1.5% (Figure 2E). The generated binary CBT and FX ROIs were manually inspected and refined as needed (by E.D.). Additionally, global WM masks were generated from the T1w-MPRAGE image using FreeSurfer (50), and then co-registered to the T1 AC-PC space. The overlap between each binary WM ROI (bilateral CBT and FX, and the global WM) and the spatial coverage of the diffusion scan (two diffusion scans for the older subject) was used to define the final WM ROIs for statistical analysis.

#### 2.3.5 Statistical analysis

All statistical analyses (Figure 2F) were conducted using the *Statistics and Machine Learning Toolbox* in MATLAB R2024b (Mathworks, Natick, MA). The mean value of each quantitative diffusion metric was first computed for each WM ROI. To assess the scan-rescan reproducibility, each ROI-averaged metric was compared between the two scans of the older group (older-1 vs. older-2), while the younger group was scanned only once. Reproducibility was evaluated using the intraclass correlation coefficient (ICC) and Bland-Altman analyses. A two-way random-effects, single-measure, absolute-agreement ICC analysis was performed, and ICC values and their 95% confidence intervals (CIs) were calculated. For Bland-Altman analysis, to account for scale differences across metrics, the relative limits of agreement (LoA_rel_) were calculated as LoA_rel_ = bias_rel_ ± 1.96×SD_rel_, where bias_rel_ and SD_rel_ were the mean and standard deviation of the relative scan-rescan difference: Δ_rel, scan-rescan_=2 × (scan 2 - scan 1) / (scan 2 + scan 1) × 100%. Subsequently, each ROI-averaged diffusion measure was averaged between the two visits for each older subject (“older-average”), and the group differences of each diffusion metric between “older-average” and younger subjects were evaluated. Given the small sample sizes (*N*=11 and 7 for the older and younger groups, respectively), data normality was first assessed using quantile-quantile (Q-Q) plots. Outliers were observed in the Q-Q plots of several metrics and WM ROIs, particularly for MK_I_ in the younger group (Supplementary Figure S1). As such, a nonparametric Mann-Whitney U test was used to evaluate unpaired age-related group differences, before (*p*_uncorr_) and after (*p*_corr_) FDR correction for multiple metrics and ROIs. Additionally, the median, 25^th^ (Q1), and 75^th^ (Q3) percentiles of each metric were calculated for the younger and older groups. The percentage differences (Δ_%_) in median values of each metric between the younger and older groups were also calculated as Δ_%_ = 2 × (median_older-average_ - median_younger_) / (median_older-average_ + median_younger_) × 100%.

## 3 Results

Quantitative diffusion parameter maps are obtained using the developed TDE protocol, with representative maps from the two scans of an older subject shown in Figure 3. The maps have consistent image quality between the two scans, with the mean scan-rescan differences across the overlapped brain parenchyma indicated on each map, ranging from -0.027 (for MK_I_) to 0.013 (for MK_A_).

**Figure 3:**
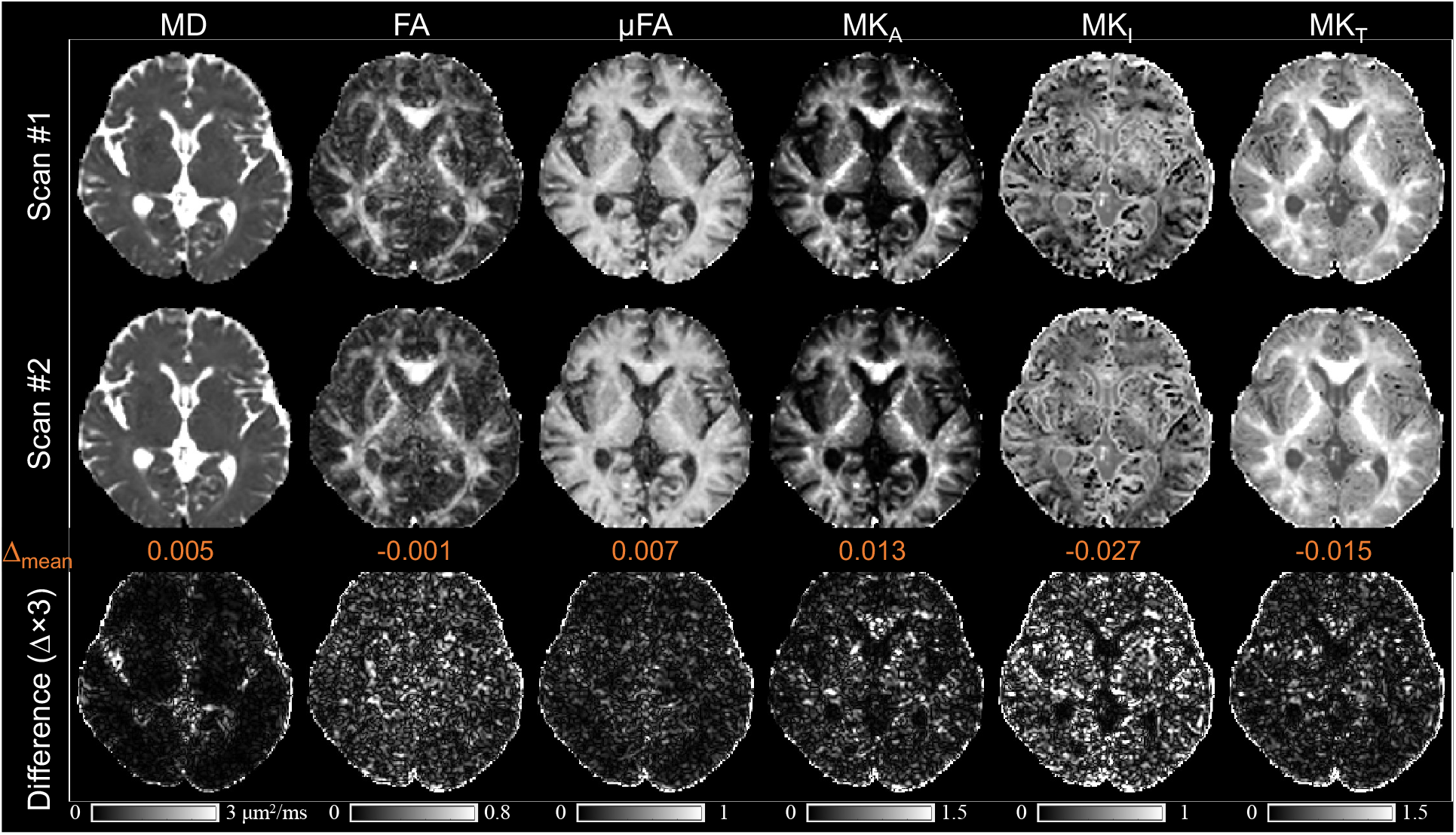
Example diffusion parameter maps of one older subject, from (**A**) scans 1, (**B**) scan 2, and (**C**) the inter-scan absolute differences (Δ×3). The mean inter-scan differences (Δ_mean_) over the brain parenchyma are marked for each metric. Abbreviations: MD: mean diffusivity; FA: fractional anisotropy; μFA: microscopic fractional anisotropy; MK_A_: anisotropic mean kurtosis; MK_I_: isotropic mean kurtosis; MK_T_: total mean kurtosis (MK_A_ + MK_I_).

The scan-rescan measurements of diffusion metrics for the older subjects across WM ROIs are plotted in Figure 4, including bilateral CBT and FX, and the global WM. The ICC and 95% CI for each diffusion metric and WM ROI are summarized in Table 1. Diffusion metrics other than MK_I_ demonstrate moderate to excellent reproducibility across all WM ROIs (ICCs≥0.64). MK_I_ shows moderate reproducibility in the global WM (ICC=0.70, 95% CI: 0.21 to 0.91) and right FX (ICC=0.69, 95% CI: 0.22 to 0.90), but shows poor reproducibility in other WM ROIs (ICC≤0.43).

**Figure 4:**
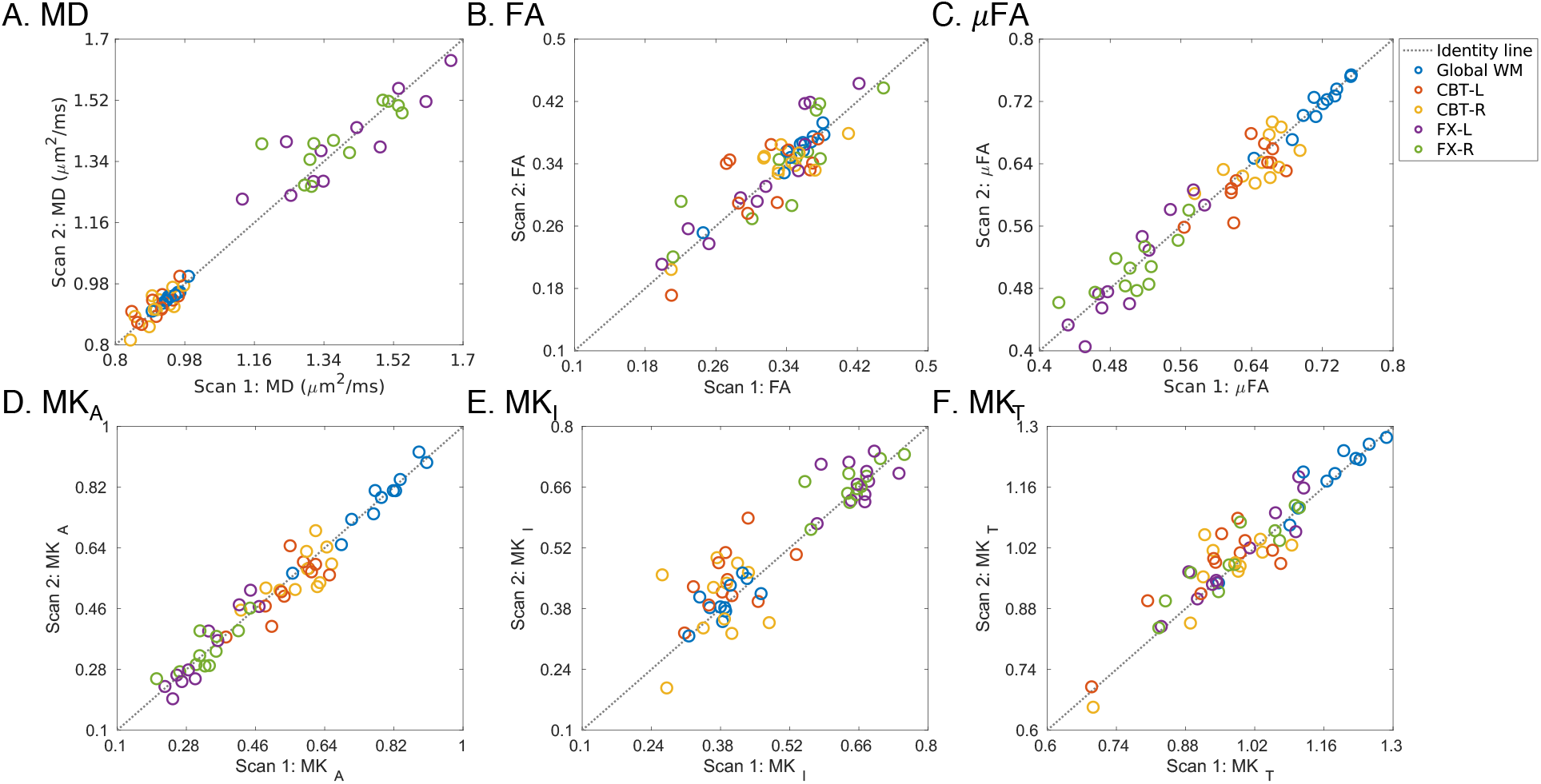
Scatter plots of scan-rescan measurements of diffusion metrics (**A**: MD, **B**: FA, **C**: μFA, **D**: MK_A_, **E**: MK_I_, and **F**: MK_T_) for the older subjects across white matter ROIs. In each plot, different ROIs are denoted by different marker colors. The line of identity is shown as a gray dotted line as a reference. The ROI-specific strength of scan-rescan reproducibility is quantified using the intraclass correlation coefficient (ICC), as summarized in Table 1. Abbreviations: MD: mean diffusivity; FA: fractional anisotropy; μFA: microscopic fractional anisotropy; MK_A_: anisotropic mean kurtosis; MK_I_: isotropic mean kurtosis; MK_T_: total mean kurtosis (MK_A_ + MK_I_); WM: white matter; CBT: temporal part of the cingulum bundle; FX: fornix; L: left side; R: right side.

**Table 1:**
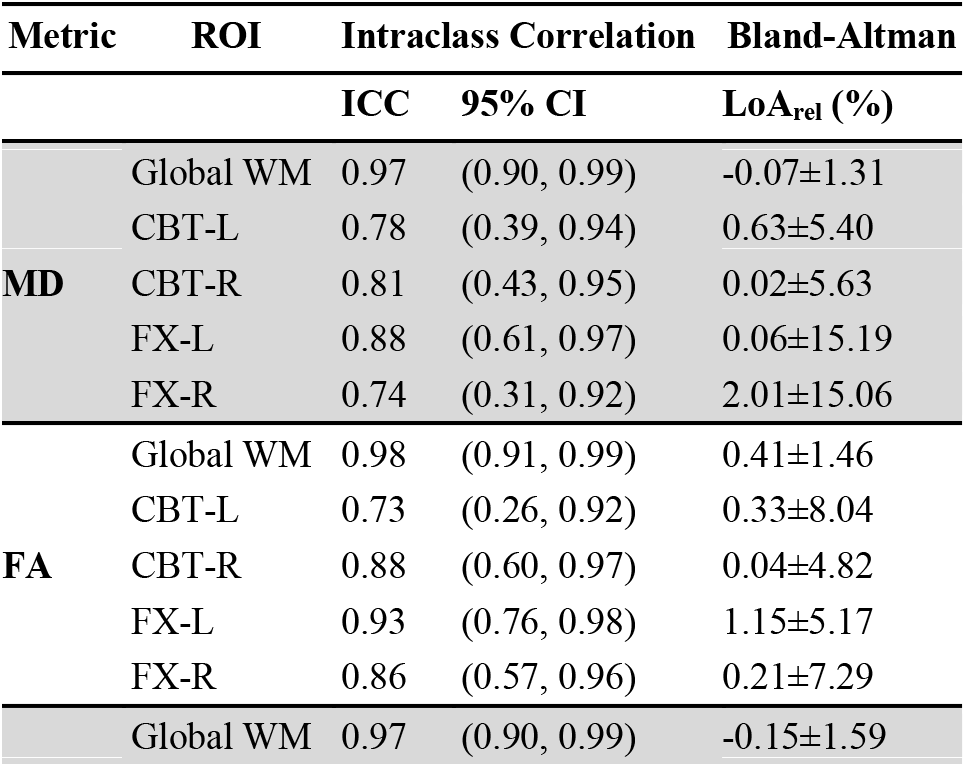
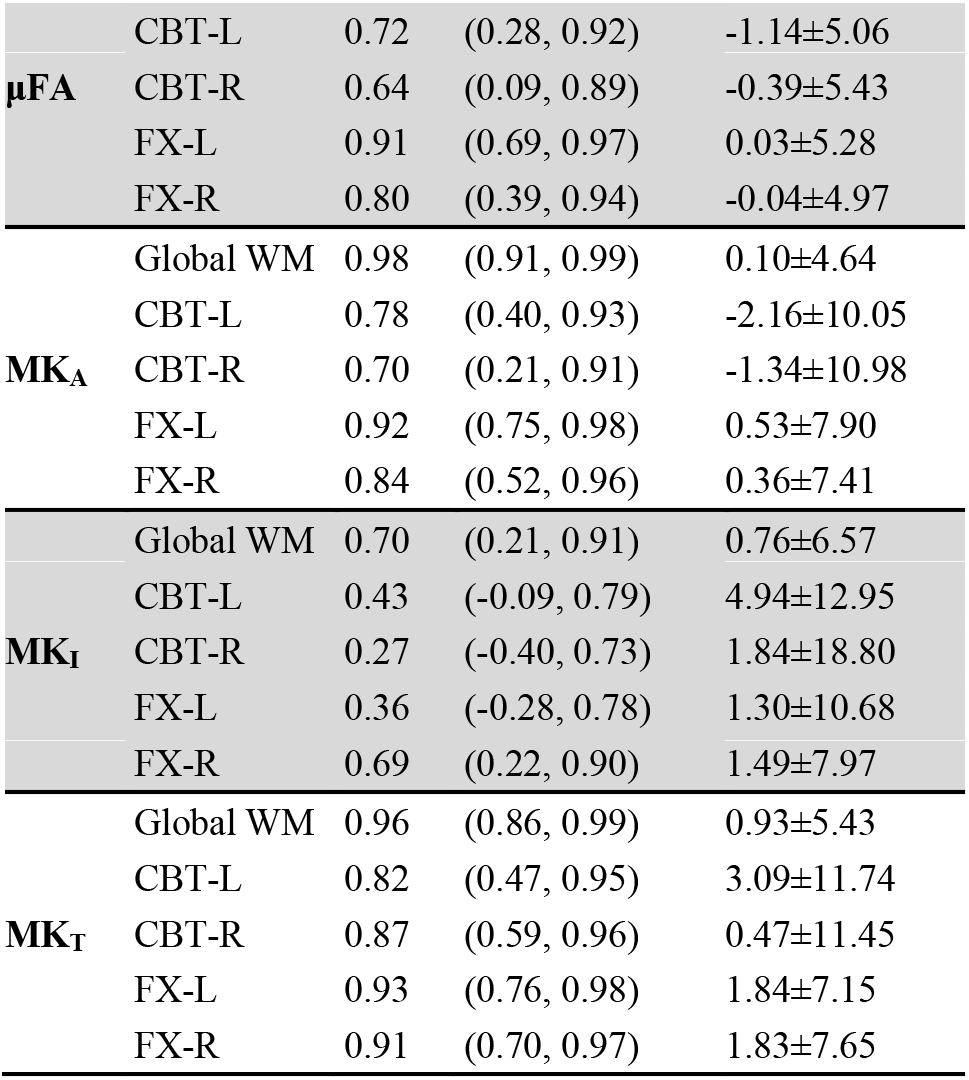
Summary of scan-rescan reproducibility analyses of the developed TDE protocol in the older cohort across white matter (WM) regions of interest (ROIs). Column 1 lists the diffusion metric names; Column 2 lists the ROI names; Columns 3 and 4 report intraclass correlation coefficient (ICC) analysis, including ICC and 95% confidence interval (CI); Column 5 summarizes the Bland-Altman analysis, using the relative limits of agreement (LoA_rel_), defined as bias_rel_ ± 1.96×SD_rel_, where bias_rel_ and SD_rel_ are the mean and standard deviation of the relative scan-rescan differences. Other abbreviations: MD: mean diffusivity; FA: fractional anisotropy; μFA: microscopic FA; MK_A_: anisotropic mean kurtosis; MK_I_: isotropic mean kurtosis; MK_T_: total mean kurtosis; CBT: temporal part of the cingulum bundle; FX: fornix; L: left side; R: right side.

The Bland-Altman plots of the scan-rescan reproducibility of diffusion metrics are shown in Figure 5, further confirming the good scan-rescan agreement. Note that the LoAs in Figure 5 (red and dark dashed lines) are derived from pooled measurements across all ROIs, while ROI-specific relative LoAs are provided in Table 1. The largest relative biases (bias_rel_≥2%) are present in MD in the right FX (2.01%), MK_A_ (-2.16%), MK_I_ (4.94%), and MK_T_ (3.09%) in the left CBT, while for other diffusion metrics and WM ROIs, biases range from -1.34% to 1.84%. The largest LoA_rel_ widths (≥20%), defined as 2×1.96×SD_rel_, are present in MD in the left (30.38%) and right (30.12%) FX, MK_A_ in the left (20.10%) and right (21.96%) CBT, MK_I_ in the left (25.90%) and right (37.60%) CBT, and in the left FX (21.36%), and MK_T_ in the left (23.48%) and right (22.90%) CBT. For each diffusion metric, no evidence of proportional bias is observed, and differences were evenly distributed across the measurement range.

**Figure 5:**
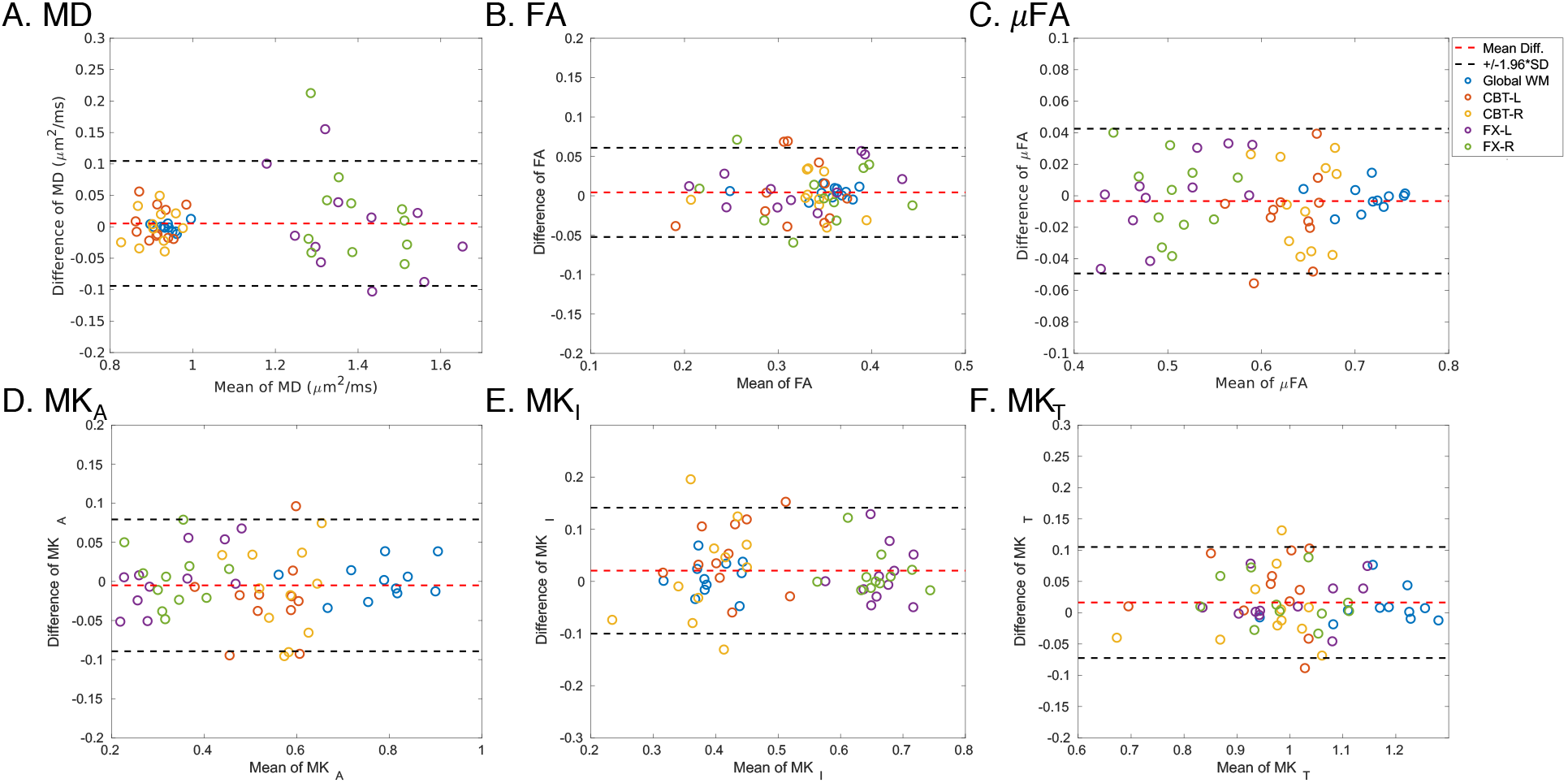
Bland-Altman analyses of the scan-rescan reproducibility of diffusion metrics (**A**: MD, **B**: FA, **C**: μFA, **D**: MK_A_, **E**: MK_I_, and **F**: MK_T_) for the older subjects across white matter ROIs. In each plot, different ROIs are denoted by different marker colors. The mean difference (bias) is shown as a red dashed line, with upper and lower limits of agreement (LoA, defined as bias ± 1.96 × SD of the scan-rescan differences) indicated by dark dashed lines. The LoAs presented in the plot are derived from pooled measurements across all ROIs, while ROI-specific LoAs are provided in Table 1. Abbreviations: MD: mean diffusivity; FA: fractional anisotropy; μFA: microscopic fractional anisotropy; MK_A_: anisotropic mean kurtosis; MK_I_: isotropic mean kurtosis; MK_T_: total mean kurtosis (MK_A_ + MK_I_); WM: white matter; CBT: temporal part of the cingulum bundle; FX: fornix; L: left side; R: right side.

The comparison results of each ROI-averaged diffusion metric between the younger and older groups across WM ROIs are plotted in Figure 6. The medians of each ROI-averaged diffusion metric in the younger and older groups, the percentage of the age-based difference of medians (Δ_%_), and *p*-values are summarized in Table 2. The most pronounced differences between the young and older groups include FA in the right CBT (8.36%) and left FX (-23.27%), uFA in the left (-18.25%) and right FX (-8.60%), MK_A_ in left (-46.93%) and right FX (-21.74%), all with *p*_uncorr_<0.05 before the FDR correction, but the differences are not statistically significant after the FDR correction.

**Figure 6:**
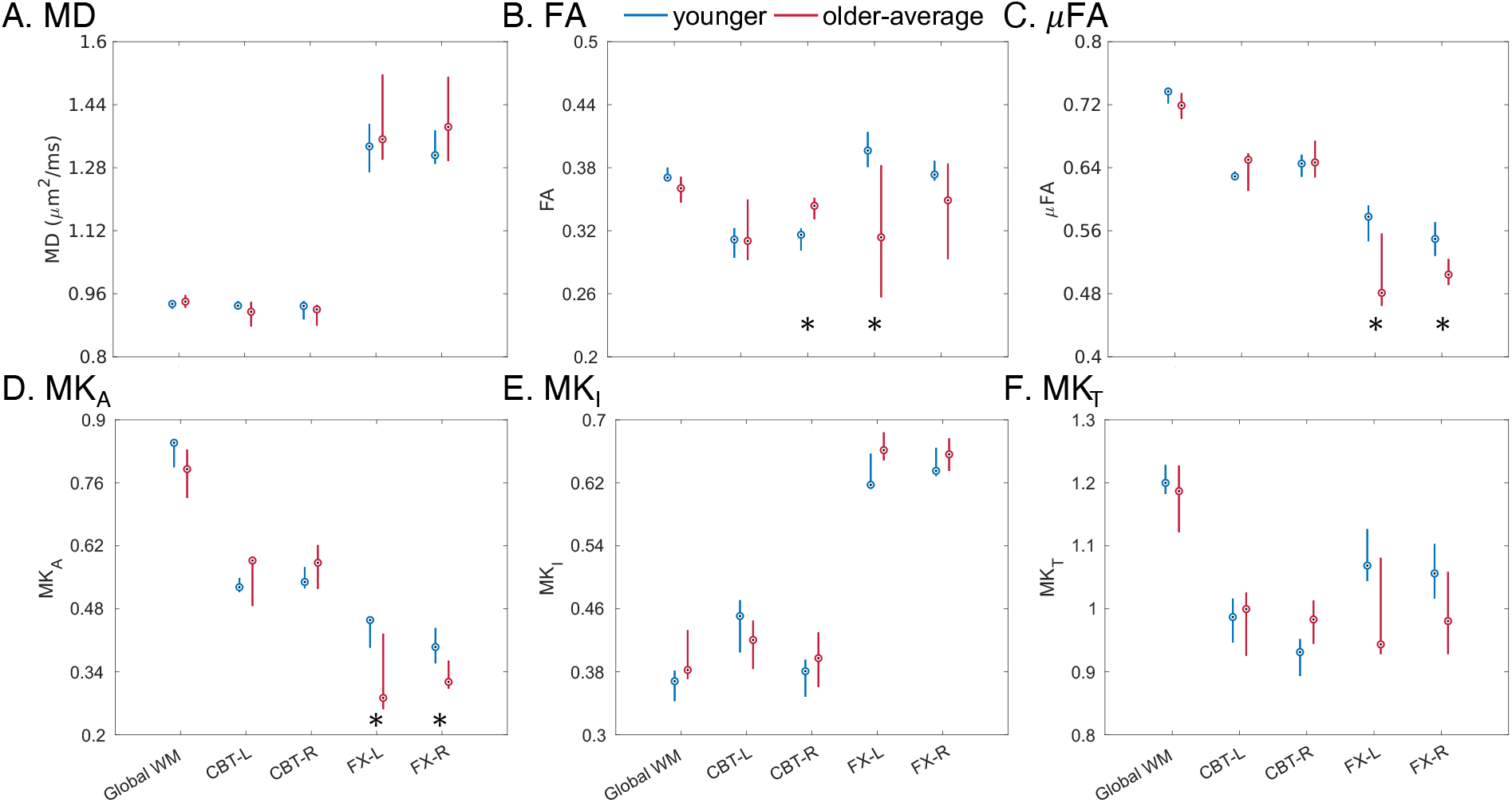
Boxplots of diffusion metrics (**A**: MD, **B**: FA, **C**: μFA, **D**: MK_A_, **E**: MK_I_, and **F**: MK_T_) between the younger (color: blue) and older (color: red) groups across white matter ROIs. For the older group, measurements are averaged across the two scans (referred to as “older-average”). In each box, the central circles denote the median values, while the down and up whiskers denote the 25th and 75th percentiles, respectively. No inter-group differences are statistically significant after the FDR correction and the inter-group differences with statistical significance before the FDR correction (*p*_uncorr_<0.05) are marked with asterisks. Abbreviations: MD: mean diffusivity; FA: fractional anisotropy; μFA: microscopic fractional anisotropy; MK_A_: anisotropic mean kurtosis; MK_I_: isotropic mean kurtosis; MK_T_: total mean kurtosis (MK_A_ + MK_I_); WM: white matter; CBT: temporal part of the cingulum bundle; FX: fornix; L: left side; R: right side; FDR: false discovery rate.

**Table 2:**
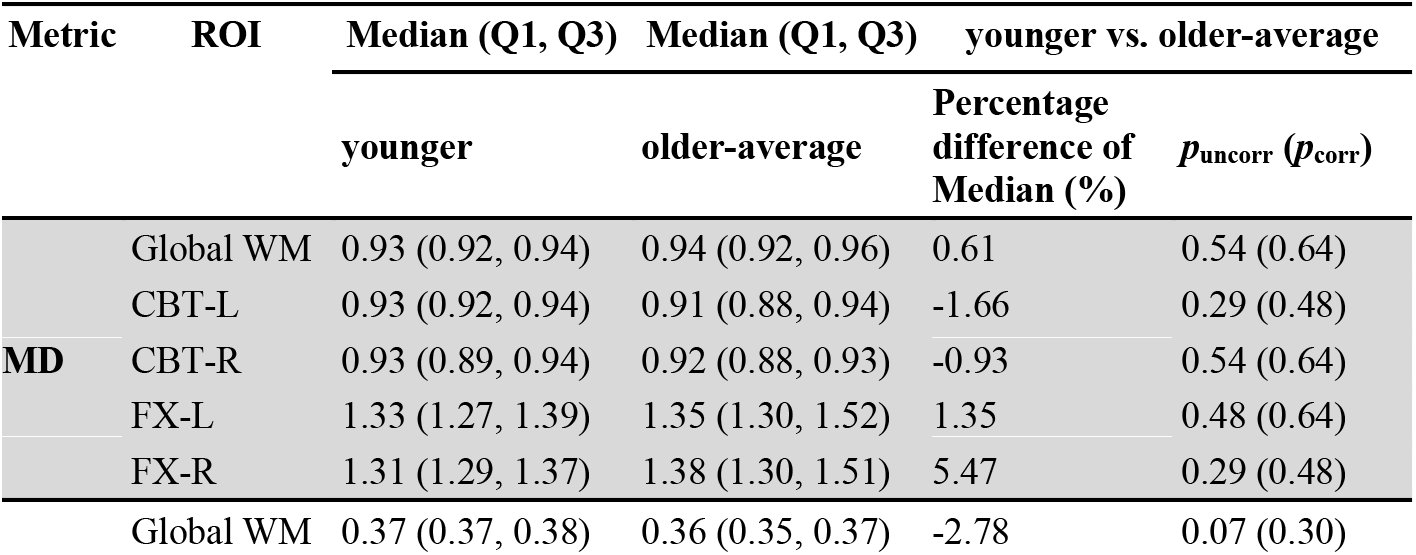
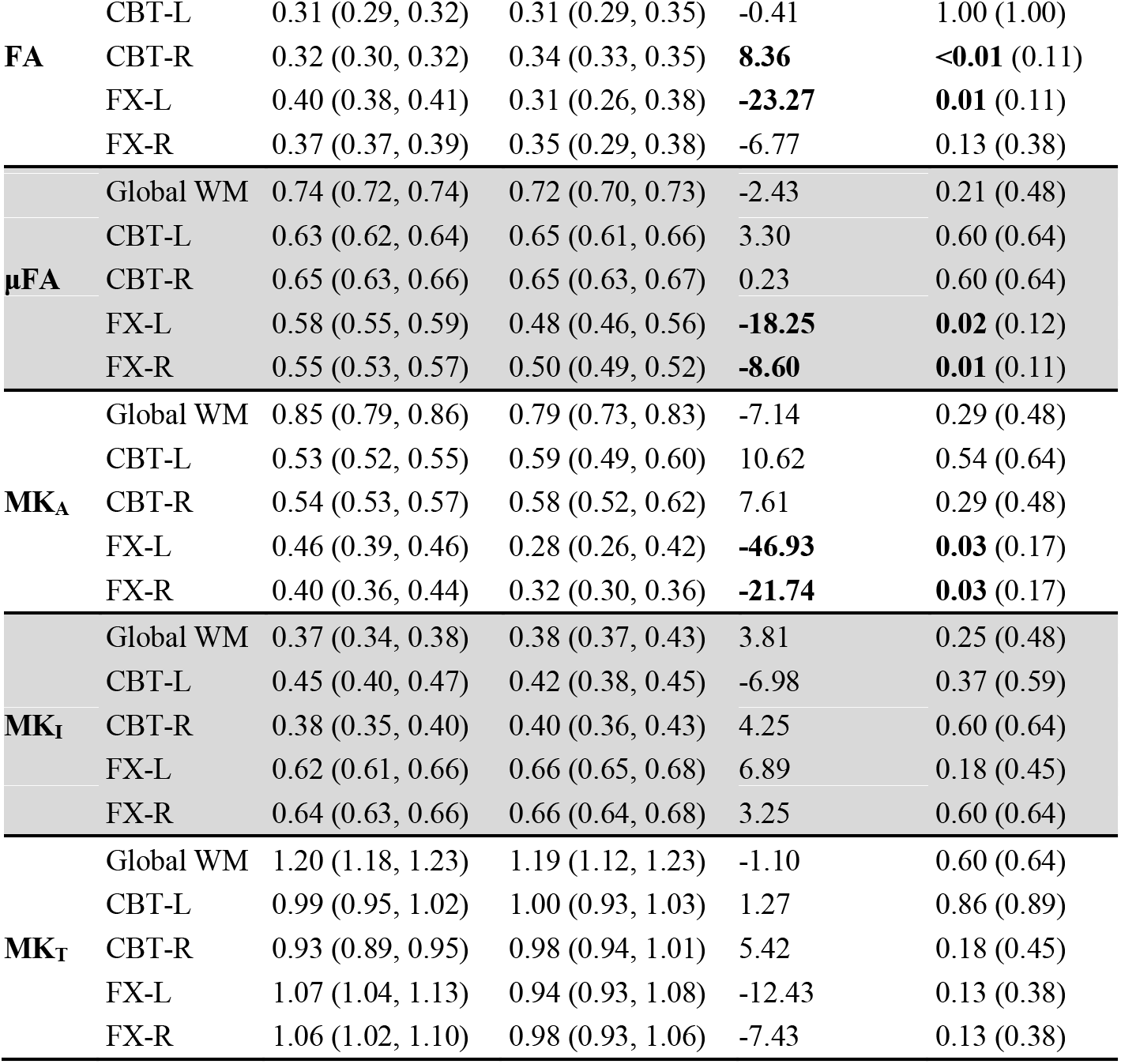
Statistical analysis of diffusion measure differences between the younger and older groups across white matter (WM) regions of interest (ROIs). For each older subject, measurements are averaged across the two scans (referred to as “older-average”). Column 1 lists the diffusion metric names; Column 2 lists the ROI names; Columns 3 and 4 report the median values along with the 25th (Q1) and 75th (Q3) percentiles for each group. Column 5 presents the percentage differences of the median values of the younger and older groups (Δ_%_). Column 6 reports *p*-values for pairwise group comparisons, before (***p*_uncorr_**) and after (***p*_corr_**) the false discovery rate (FDR) correction. *P*-values<0.05 and corresponding Δ_%_ values are highlighted in bold. Other abbreviations: MD: mean diffusivity; FA: fractional anisotropy; μFA: microscopic FA; MK_A_: anisotropic mean kurtosis; MK_I_: isotropic mean kurtosis; MK_T_: total mean kurtosis; CBT: temporal part of the cingulum bundle; FX: fornix; L: left side; R: right side.

## 4 Discussion

The principal contribution of this work is the development of an high-resolution TDE for imaging the MTL within a clinically feasible acquisition time by jointly using (1) an optimized STE waveform to compensate for the decreased diffusion encoding efficiency of STE, (2) an SNR-efficient MB-MS readout and reconstruction, and (3) a strong gradient coil to shorten the diffusion encoding duration and readout window. Collectively, a TDE protocol with an isotropic resolution of 1.8 mm, a brain coverage of ∼70 mm, and a clinically feasible scan time of ∼12 minutes has been developed. Our preliminary results have demonstrated high image quality and good scan-rescan reproducibility in the SNR-challenging MTL region, including bilateral CBT and FX. Exploratory comparisons show age-related differences in FA, μFA, and MK_A_ with *p*<0.05 before FDR correction, particularly in the fornix, indicating their potential for mapping brain microstructural changes in the MTL during neurodegeneration.

The CBT and FX in the MTL are selected as the target WM bundles because of their critical roles in cognitive function, such as memory, and in early AD progression, such as tau propagation (22,23). Notably, the most pronounced age-related differences in diffusion metrics were observed primarily in the FX in this study (Table 2 and Figure 6), further underscoring the importance of characterizing FX microstructural changes during normal and pathological aging. However, imaging the FX is non-trivial due to its small size, deep-brain location, and susceptibility to partial volume effects from cerebrospinal fluid (CSF) (24), all of which necessitate high-resolution diffusion MRI. As such, this study aims to optimize a TDE protocol of high resolution and high SNR for studying microstructural changes in neurodegeneration, within a clinically feasible time. We primarily focus on evaluating the performance of the final optimized protocol, including the scan-rescan reproducibility (particularly in the older cohort), and the sensitivity to microstructural differences between the younger and older groups. In contrast, no comparisons were performed between the final optimized protocol and any intermediate optimized protocols. The influence of each optimization step on the SNR improvement has been reported in related literature (17,25,36). In addition to synergizing among different SNR optimization strategies, the trade-off between spatial and q-space resolution should be carefully balanced with the consideration of the available hardware capabilities and total scan time. For example, a previous study has reported a specific application of super-resolution with rotating-view multi-slab acquisition in STE to achieve 1.6 mm isotropic high resolution with b-values up to *b*_max_=4 ms/um^2^ (32). However, the implementation of super-resolution can substantially increase the acquisition time of each diffusion volume (33.6 s), leaving limited space for deploying q-space sampling within a clinically feasible scan time. In our study, using an optimized 2D MB-MS readout, we achieved an isotropic resolution of 1.8 mm at *b*_max_=2 ms/um^2^, with an acquisition time of 6.6 s for each diffusion volume.

The developed TDE protocol shows a good scan-rescan reproducibility (ICCs≥0.64) for all diffusion metrics except MK_I_, across all WM ROIs. The larger scan-rescan deviations of MK_I_ were consistently reported in a previous TDE study (51). Moreover, when comparing the Bland-Altman analysis results of diffusion metrics in the global ROI between our study and Ref. (51), our study shows narrower LoAs than Ref. (51) for including MD, uFA, MK_A_, and MK_I_. This improvement may be attributed to the improvement of SNR and resolution in this study.

Among all diffusion metrics and WM ROIs, FA in the right CBT and left FX, uFA in the bilateral FX, and MK_A_ in the bilateral FX show statistically significant age-related differences before the FDR correction (Table 2). Given the small sample size, however, these findings should be interpreted with caution, as the limited statistical power may affect the robustness of the observed group differences. For example, although FA in the right CBT shows *p*_uncorr_<0.01, its increase from the younger to older group is contrary to findings reported in previous studies (4,52,53), warranting further evaluation in a larger cohort. FA in the left FX, uFA in the left FX, and MK_A_ in the bilateral FX exhibit both relatively large percentage differences between the younger and older groups (|Δ_%_|≥18.25%) and *p*_uncorr_<0.05, suggesting potentially more plausible changes in macroscopic and microscopic anisotropies during the aging process. Additional trends are observed across the diffusion metrics despite the absence of statistical significance. MD and MK_I_ are similar between the global WM and CBT but are lower than those in the FX, suggesting higher diffusivity and greater diffusional heterogeneity in the FX. Differences between FA and μFA are also observed across all WM ROIs, indicating the presence of fiber orientation dispersion for them. Overall, given the limited sample size, these age-related analyses should be considered a proof-of-concept demonstration of the optimized TDE protocol rather than a definitive characterization of aging-related microstructural changes. Future studies in larger cohorts, including patients with neurodegenerative diseases, are warranted to establish the sensitivity and clinical utility of the proposed TDE protocol for detecting microstructural changes associated with aging and neurodegeneration.

The TDE protocol can be further optimized in several aspects. First, CSF suppression has been reported to mitigate the partial volume effects on imaging the FX and improve the accuracy of DTI measures in the FX (8,24), which can also be combined with the developed TDE protocol. Second, a relatively dense q-space sampling is currently used for both LTE and STE. Further scan acceleration can be achieved by optimizing the q-space sampling and thus reducing the number of samplings (21), which will further facilitate the clinical transferability of the TDE protocol. Third, as a proof of concept, a partial brain coverage of 70 mm is tested to cover the MTL region for all the subjects in this study. In the future, the saved scan time by optimizing the q-space sampling can also be converted to a larger FOV, ideally covering the whole brain. Whole-brain coverage will enable the study of white matter integrity and connectivity changes across a broader spatial scale throughout the course of AD. Fourth, the UHP gradient used in this study is still a whole-body gradient coil, and the actual G_max_ and SR_max_ performances are more constrained by peripheral nerve stimulation (PNS) and gradient heating, compared with emerging head-only gradient coils. Combining the SNR-optimized diffusion encoding and readout with high-performance head-only gradient coils can offer the opportunity to further advance the achievable resolution and SNR, within a clinically feasible scan time (54). Lastly, the current technical evaluation focuses on WM, including the CBT and FX, two WM bundles closely associated with the early propagation of tau. The developed TDE protocol can also be used to study early microstructural alterations in gray matter, such as the entorhinal cortex and hippocampal subfields, both of which are closely associated with the origin and early propagation of tau pathology (55,56).

## 5 Conclusion

In summary, an SNR-efficient TDE protocol with an isotropic resolution of 1.8 mm, a brain coverage of ∼70 mm, and a clinically feasible scan time of ∼12 min has been developed and demonstrated to provide reproducible quantitative assessment of white matter microstructure in the MTL. This high-resolution TDE protocol provides a practical foundation for future studies of microstructural changes associated with aging and neurodegenerative disease.

## Data Availability

Data produced in the present study are available upon reasonable request to the authors

## 6 Acknowledgments

This work was supported by NIH grants K99AG080076, R01NS095985, and R01NS125781. The numerical optimization toolbox for the gradient waveform (NOW) can be found at https://github.com/jsjol/NOW. The DESIGNER preprocessing toolbox can be found at https://github.com/NYU-DiffusionMRI/DESIGNER-v2. The multi-band, multi-shot diffusion MRI reconstruction with the joint use of structured low-rank constraints and explicit phase mapping (JULEP) can be found at https://github.com/daiep/JULEP.

## 7 Author Contributions

Erpeng Dai: Conceptualization, Methodology, Investigation, Data Curation, Formal Analysis, Visualization, Writing - Original Draft Preparation, Funding Acquisition.

Xuetong Zhou: Investigation, Data Curation, Software.

S Shailja: Investigation, Data Curation, Writing - Review & Editing.

Martin K Schneider: Investigation, Data Curation.

Molly A Millar: Investigation, Data Curation.

Michael Zeineh: Conceptualization, Methodology, Formal Analysis, Writing - Review & Editing.

Carl-Fredrik Westin: Methodology, Supervision, Writing - Review & Editing, Funding Acquisition.

Jennifer A McNab: Conceptualization, Methodology, Supervision, Writing - Review & Editing, Funding Acquisition.

## 9 Data and code availability statements

Anonymized data are available on reasonable request to the corresponding author from qualified investigators.

## 10 Declaration of the use of AI

With respect to the research described in the paper: No AI tools were used in planning and carrying out the research.

With respect to writing the paper: We used GPT-5.6 to check spelling and grammar.

## 11 Supplementary Materials

**Supplementary Figure S1:**
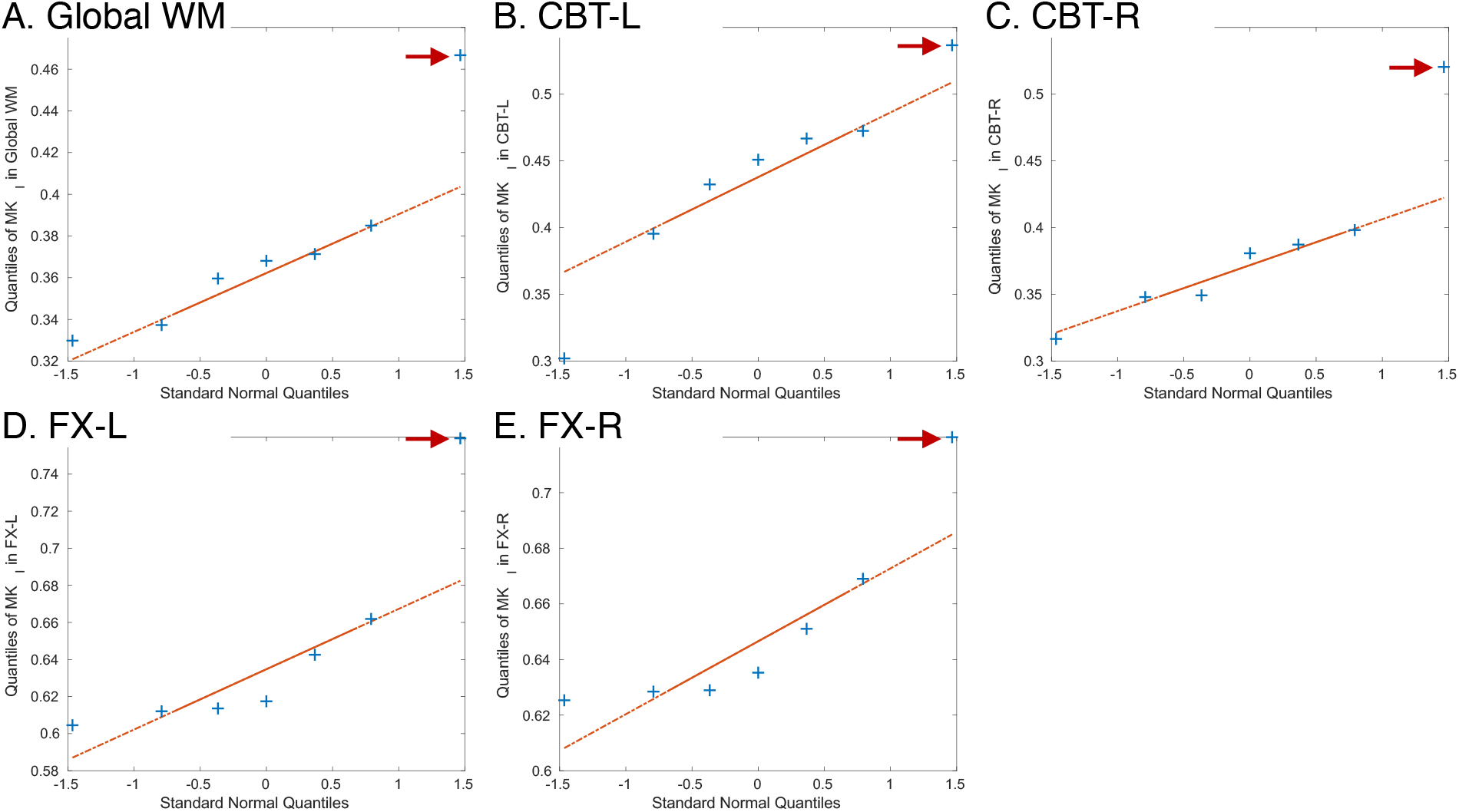
Quantile-quantile (Q-Q) plots of MK_I_ measures in different white matter (WM) ROIs (**A**: global WM, **B**: CBT-L, **C**: CBT-R, **D**: FX-L, and **E**: FX-R), in the younger group. Outliers are observed in each plot (indicated with red arrowheads). Abbreviations: MK_I_: isotropic mean kurtosis; CBT: temporal part of the cingulum bundle; FX: fornix; L: left side; R: right side.

